# HEALTH PROMOTION AND PREVENTIVE INTERVENTIONS FOR SENIOR CITIZENS IN COUNTRIES WITH ACCESSIBLE AND FUNDED HEALTH CARE: STUDY PROTOCOL FOR A SCOPING REVIEW

**DOI:** 10.1101/2024.02.08.24302500

**Authors:** Petra Vikman Lostelius, Henri Aromaa, Malin Lohela Karlsson, Åsa Revenäs, Lena Nordgren

**Author notes:** Corresponding author (LN).

## Abstract

To improve health in a growing population with increasing life expectancy, a stepwise health care program for citizens aged 65 years and older will be implemented in Region Västmanland in Central Sweden. This scoping review builds on a previous review and aims to map interventions in a larger number of countries and from 2019 to present. The objective of this scoping review is to map the literature on the components of health promotion and preventive interventions in countries that provide accessible and funded health care. The JBI methodology will be employed for good structure. Searches will be conducted in databases and websites, for grey literature. The searches will include health promotion studies, preventive health measures, intervention methods, intervention delivery, outcome measures, or health-economic evaluations for people aged 65 years and older. Two reviewers will individually assess each study and extract data. Data analysis will consist of basic frequencies and percentages. The results will be presented in tables and/or diagrams and a narrative summary.

## Introduction

The primary objective of this scoping review is to systematically map the existing literature, providing a comprehensive overview of health promotion and preventive interventions for older individuals in countries that offer accessible and funded healthcare for their citizens. The review aims to identify and present detailed information on the providers of health promotion and prevention programs, elucidating the components included in preventing diseases and promoting health. Additionally, it seeks to outline the outcome measures employed to identify health risks, the modes of intervention delivery, and the assessment of intervention effects.

Furthermore, the scoping review will investigate which outcome measures are commonly utilized, assess the effects of interventions, and consider any health economic evaluations conducted. Another important aim is to pinpoint current knowledge gaps in the realm of health promotion and preventive interventions for older individuals in countries with accessible and funded healthcare systems. By addressing these objectives, the review endeavours to contribute to a better understanding of the landscape of healthcare interventions for the elderly, identifying areas that may require further research and development.

### Health and health interventions for older people

The ageing world population is well known. As the population grows older, the demands on health care increase, which implies the need for health care to adjust. The World Health Organization’s (WHO) Agenda 2030 for Sustainable Development contains, among others, the goal of ensuring good health and the promotion of wellbeing for all at all ages [1]. An increasing life expectancy leads to an older and larger population in which the proportion of people with reduced health increases due to one or more chronic diseases [2]. Health in the Swedish population is overall good [3], with a life expectancy age of approximately 85 years for women and 81 years for men at present in 2023 [4]. In 2021, approximately 25% of people older than 80 years were admitted for hospital care at least once [5].

To support the health and quality of life of older people, health promotion and health prevention incentives are important. Health promotion can be defined as a process that enables improved control of life and health, while disease prevention is an action that limits or hinders the development of disease [6,7]. Health promotion should be provided in all contexts, such as educational settings, workplaces, and households (6), to avoid disease or traumatic incidents. This requires multidimensional knowledge, for example, about physical activity and social aspects of human needs. The promotive approach involves cooperation between society and individuals and includes the ambition to identify and strengthen a person’s resources, participation and responsibility for a healthy life [8]. Health promotion builds on the knowledge of what helps people evolve and thrive.

Preventive interventions in health care involve knowledge about what creates poor health and disease [8] and include screenings, check-ups, and patient counselling to prevent illnesses, disease, or other health problems. Prevention of the ageing population in society is necessary and should focus on interventions that are evidence-based, such as the use of medications, personalized health management, fall prevention and vaccination [9]. Lifestyle issues, for example, smoking status, physical activity level, nutrition, and functions such as vision, hearing, and dentition, along with screenings for noncommunicable diseases [10], such as hypertension, hyperlipidaemia, osteoporosis, cardiovascular disease, and several cancer types, are important for investigating senior citizens and should certainly include the patient in discussions and decisions to the greatest extent possible [11]. Preventive health work should focus not only on mortality but also on function and health-related quality of life [10].

From approximately 65 years of age, an increasing number of people are at risk of developing frailty [12]. Frailty is a multidimensional condition that includes vulnerability to biopsychosocial factors [13]; thus, an integral conceptual model and a multidimensional approach are needed [13,14]. Frailty is a condition of accelerated biological ageing that causes a loss of ability to adapt to physical, mental and social stressors [15] and thus affects a person’s ability to manage it every day. For example, frail people have a higher risk of falling, becoming increasingly dependent on others, having an increased risk of being admitted to the hospital, becoming institutionalized, and having an increased risk of dying [15,16].

To inform decision-makers, research councils and researchers, a scoping review investigated health prevention in community-dwelling old people living in Nordic countries (Sweden, Norway, Denmark, Iceland, Finland, and the Faroe Islands) [17]. The included interventions could be single- or multicomponent and included fall prevention programs, general health promotion, health promotion with a focus on physical activity, and disability prevention interventions. Interventions were excluded if they were directed towards a specific disease or diagnosis, dental health, older people’s cognitive malfunctions, or programs assessing medication effects or focused on specific body structures. Furthermore, only randomized controlled studies were included. The review concluded that although the magnitudes of effect and the number of follow-ups differed, there were overall improved health outcomes. Senior meetings, preventive home visits and exercise interventions were recommended for community-dwelling older people. However, to provide a solid ground for decision-makers on implementation of health promotion or prevention interventions, only health outcomes do not provide enough information. The review revealed a lack of studies related to cost-effectiveness, the experiences of participants and the feasibility of the interventions.

### Study rationale

A scoping review will be performed that builds on the previously mentioned scoping review by Bajraktari et al. (12); i.e., the population and review questions will be the same, but the literature search will cover a more recent time span and include more countries and articles with all study designs. The review will add to the available literature from 2019 to the present in countries with health care systems that strive to ensure universal access to health care for all citizens through publicly funded or regulated systems.

### Overall study objective

To map the literature on the characteristics of health promotion and preventive interventions for older people, 65 years and older, in countries that provide accessible and funded health care for their citizens.

### Overall research question

What health promotion and preventive interventions have been used (concept) for people aged 65 years and older (population) in countries with accessible and publicly funded health care systems (context)?

### Review questions

1. In which contexts have the interventions been conducted?
2. For which populations, from 65 years and older, have interventions been conducted?
3. How have the interventions been designed (e.g., which components, duration of interventions and mode of delivery)?
4. Which feasibility aspects have been described?
5. How have the participants experienced the interventions?
6. Were the interventions effective, and on which outcomes?
7. Were interventions cost-effective?

## Materials and methods

The scoping review will follow the JBI approach for scoping reviews [18,19] and utilize the PRISMA-ScR reporting guideline and checklist [20]. This decreases the risk for bias from arbitrary decisions, duplication of existing reviews, and the detection of selective reporting easier to identify [21]. This process includes the involvement of two or more researchers and an a priori review protocol. In case of any deviation from the study protocol, notes will be taken, and alterations will be accounted for in the review manuscript to ensure consistency and reproducibility [19]. The review questions were formed in an iterative process among the SHIELD members (PVL, LN, MLK and ÅR) using the population content context (PCC) framework, which states the population, concept, and context [19].

### Inclusion criteria

#### Population

Eligible studies will provide at least one abstract in English and include people aged 65 years without known previous diagnoses.

#### Concept

The concept of interest is interventions that promote health and prevent disease. The scoping review aims for an extensive overview of the interventions and will thus include single-component and multiple-component promotive or preventive intervention programs.

#### Context

All health promotion and prevention incentives in health care, municipality, community, and social settings in countries with health care in an accessible and publicly funded health care system; Norway, Denmark, Finland, Iceland, Canada, Netherlands, United Kingdom, Australia, New Zealand, France, Germany, Belgium, Austria, Italy, Spain, Portugal, Switzerland, and Sweden. No exclusion will be made for not being delivered within the health care system.

### Exclusion criteria

To ensure the current literature and to include healthy individuals, the exclusion criteria are articles written before 2019, studies in populations with extensive need for support in daily activities, studies with health interventions that focus on a specific disease or diagnosis, dental health, older people’s cognitive malfunctions, or interventions assessing medication effects or focusing on specific body structures.

### Types of evidence sources

The scoping review will include all the research methodologies and unpublished (grey) literature.

#### Search strategy

Evidence sources in English published from 2019 to December 31, 2023, will be considered. The timeframe was chosen to identify relevant evidence on the topic. A research librarian (HA) has participated from the start and has been offered coauthorship.

A primary literature search in PubMed to identify appropriate keywords and terms and developed the search strategy (Appendix 1) was performed by PVL and HA on November 14th, 2023. The approach to the search followed the same recommendations as the JBI systematic reviews [22].

The first step of the search strategy was to examine PubMed for relevant sources using an initial search strategy (Appendix 1). The search strategy was subsequently adapted and refined for use in PubMed, PsycInfo, Amed and Cinahl. These databases are accessible to the authors, can identify existing reviews, and allow for broad study inclusion. Protocols, policy documents, and supplementary literature were searched for using Google, Google Scholar, and ClinicalTrials.gov. Additional studies that were not previously identified were identified in the reference lists of the selected studies. The preliminary search strategy is included in Appendix A. The structured search occurred on December 15, 2023, through December 31, 2023.

#### Selection of sources of evidence

The identified records from the database and grey literature search will be subsequently screened with Rayyan, a free-of-charge artificial intelligence tool (Rayyan - AI Powered Tool for Systematic Literature Reviews for Organizing and Managing Reviews), which allows a blinded screening process.

In Rayyan, the records will be deduplicated, screened and selected in compliance with the inclusion criteria. The key text words in the title and abstract initiate the selection process. If deemed potentially relevant, an examination of the full-text article will be performed against the inclusion criteria. The reasons for the exclusion of full-text articles will be recorded and reported. The selection criteria will be mapped using the PRISMA-ScR flow chart, which shows the inclusion and exclusion process.

## Summary of the evidence

Scoping reviews are not obligated to assess the methodology or strength of evidence in the included studies [20]. This is also not necessary, considering the research questions, and will therefore not be performed.

### Data extraction

Basic descriptive data (authors, title, year published) and data consistent with the review questions and inclusion criteria will be extracted and presented in a draft table in the protocol. To assess the feasibility of the draft extraction tool for the review, a pilot test will be performed with a small sample of articles. The extraction of data will be performed independently by PVL and LN, and the extracted data will be crosschecked. Disagreements will be resolved by consulting ÅR or MLK and discussions within the group.

### Data analysis and presentation

The basic frequencies and percentages of the data will be analysed descriptively. The data will be presented in tables accompanied by a narrative summary of the extracted information.

Themes and/or diagrams will illustrate potential trends and accompany a narrative summary.

### Ethics and dissemination

No ethical approval is needed. The study results will be published in a suitable peer-reviewed scientific journal.

## Data Availability

No datasets were generated or analysed during the current study. All relevant data from this study will be made available upon study completion.

## Acknowledgements

We thank the project manager of the Senior Health Intervention Program, Region Västmanland, for the research opportunity.

## Appendix 1 Search strategy

MESH Terms Health promotion

[Title/Abstract] prevention OR intervention OR “health programme” OR “health program” OR “health education”

1 OR 2

MESH Aged

[Title/Abstract] (“old people” OR aged OR “old population” OR senior OR elderly OR “older adults” OR “well older people” OR “senior citizen” OR “old age” OR “advanced age” OR geriatric OR ageing OR aging)

4 OR 5

[Title/Abstract] (“independent living” OR “community dwelling” OR “home dwelling” OR “community living” OR “living alone” OR “ageing in place”)

3 AND 6 AND 7

Norway[MeSH Terms]) OR (Denmark[MeSH Terms])) OR (Finland[MeSH Terms])) OR (Iceland[MeSH Terms])) OR (Canada[MeSH Terms])) OR (Netherlands[MeSH Terms])) OR (United Kingdom[MeSH Terms])) OR (Australia[MeSH Terms])) OR (New Zealand[MeSH Terms])) OR (France[MeSH Terms])) OR (Germany[MeSH Terms])) OR (Belgium[MeSH Terms])) OR (Austria[MeSH Terms])) OR (Italy[MeSH Terms])) OR (Spain[MeSH Terms])) OR (Portugal[MeSH Terms])) OR (Switzerland[MeSH Terms])) OR (Sweden[MeSH Terms]))

8 AND 9

## Appendix 2 Data extraction tool

### Study details

Author

Title of publication

Year of publication

Journal name (in full)

Specific country in which the study was conducted

Study design

Study purpose

List of outcome measures in the study (if any)

Population

Age

### Intervention type

Health promotion/preventive intervention methods

Intervention context

Intervention components, duration, mode of delivery

Outcome measures

Feasibility aspects

Participants experiences of the intervention

Effectiveness of outcomes

**Health economic evaluations and their indications**

